# Differences in site-specific cancer incidence by individual- and area-level income in Canada from 2006-2015

**DOI:** 10.1101/2023.01.13.23284277

**Authors:** Parker Tope, Samantha Morais, Mariam El-Zein, Eduardo L. Franco, Talía Malagón

**Author notes:** Talía Malagón. **Correspondence:** Name: Talía Malagón, Affiliation: Division of Cancer Epidemiology, McGill University.

## Abstract

**Introduction:** Income, a component of socioeconomic status, influences cancer risk as a social determinant of health. We evaluated the independent associations between individual- and area-level income, and site-specific cancer incidence in Canada.

**Methods:** We used data from the 2006 and 2011 Canadian Census Health and Environment Cohorts, which are probabilistically linked datasets constituted by 5.9 million and 6.5 million respondents of the 2006 Canadian long-form census and 2011 National Household Survey, respectively. Individuals were linked to the Canadian Cancer Registry through 2015. Individual-level income was derived using after-tax household income adjusted for household size. Annual tax return postal codes were used to assign area-level household income quintiles to individuals for each year of follow-up. We calculated age-standardized incidence rates (ASIR) and rate ratios for cancers overall and by site. We conducted multivariable negative binomial regression to adjust these rates for other demographic and socioeconomic variables.

**Results:** Individuals of lower individual- and area-level income had higher ASIRs compared to those in the wealthiest income quintile for head and neck, oropharyngeal, esophageal, stomach, colorectal, anal, liver, pancreas, lung, cervical, and kidney and renal pelvis cancers. Conversely, individuals of wealthier individual- and area-level income had higher ASIRs for melanoma, leukemia, Hodgkin’s lymphoma, and non-Hodgkin’s lymphoma, breast, uterine, prostate, and testicular cancers. Most differences in site-specific incidence by income quintile remained after adjustment.

**Conclusions:** Although Canada’s publicly funded healthcare system provides universal coverage, inequalities in cancer incidence persist across individual- and area-level income gradients. Our estimates suggest that individual- and area-level income affect cancer incidence through independent mechanisms.

## INTRODUCTION

Socioeconomic status (SES), a well-documented health determinant, is comprised of several demographic and economic factors such as education, occupation, and income (1). The combined effect of these factors shapes an individual’s behaviours, lifestyle, built environment, and access to opportunity and healthcare which, in turn, impacts an individual’s risk of cancer and other health outcomes (1).

Income, a fundamental component of SES, can be classified as individual- and area-level, both of which provide distinct insight into the assessment of socioeconomic inequalities in health outcomes. Individual-level income captures a person’s immediate funds whereas area-level income provides contextual information about an individual’s built environment, such as community services, social and physical conditions, infrastructure quality, and access to healthcare (1). These two measures of income contextualize an individual’s habitus, or societal dispositions, arising from their personal and residential economic conditions (2). While area-level income measures are often considered in research as proxies of individual-level income, there is increasing evidence that the effect of area-level income is a distinct component of SES independent of individual-level income (1, 3, 4). Although both income measures are correlated – an individual’s income influences their area-level income and vice versa – identifying the independent effect of each on cancer incidence can help explain how protective and risk factors for cancer are differentially distributed across the SES gradient (1). Prior investigations into the impact of SES on cancer incidence in high income countries have mostly relied upon area-level income as a proxy for individual-level SES (5–8), household income unadjusted for household size (9, 10), or other individually-measured components of SES other than income (e.g., education) (11). What has yet to be established are the separate effects of an individual’s personal and neighbourhood wealth on their cancer risk, and the concordance between the two. Quantifying these effects could further clarify the utility of area-level income as a proxy for individual-level income (4).

Given that cancer remains the second leading cause of mortality worldwide (12), evaluations of cancer risk across area- and individual-level SES gradients are essential for identifying at-risk subpopulations and informing future cancer prevention efforts. Canada is a country with a publicly funded health system providing universal coverage to maintain equitable accessibility and quality of care to all residents regardless of their financial means. However, previous research has demonstrated that inequalities in cancer incidence persist across the SES gradient for particular cancer sites even with universal health coverage (13–19). These studies only evaluated area-level income as a proxy for individual-level income (13–18), or obtained individual-level data via surveys administered to populations not representative of the entire Canadian population (19). To our knowledge, none have determined the independent effects of both individual- and area-level income on cancer incidence in Canada due to difficulty linking individual-level income data with cancer data. Cancer registries, such as the Canadian Cancer Registry (CCR) in Canada, or the Surveillance, Epidemiology, and End Results (SEER) program in the United States, capture residential demographic information, but not household-level socioeconomic data, which only permits probabilistic identification of patients’ area-level income and no measure of individual-level income. Given this difficulty, research into the independent effects of individual- and area-level income, two fundamental components of SES, is needed to determine how they influence cancer risk.

In this study, we aimed to quantify the independent associations between individual- and area-level income, and overall and site-specific cancer incidence in Canada between 2006 and 2015 through data linkage of individual-level socioeconomic data to the CCR.

## METHODS

### STUDY POPULATION & DATA SOURCES

#### Study population

We used data from the 2006 and 2011 Canadian Census Health and Environment Cohorts (CanCHECs) to quantify the associations between individual- and area-level income quintile, and cancer incidence between 2006 and 2015. The 2006 and 2011 CanCHECs are population-based datasets constituted by respondents to the 2006 long-form census and 2011 National Household Survey (NHS) who could be probabilistically linked to the derived record depository (DRD), a population data file comprised of individuals living in Canada (20).

The 2006 census consisted of short- and long-form surveys sent out to usual Canadian residents, or residents who spend a majority of their time living in Canada, who were non-institutionalized, or not in a collective dwelling, such as a hospital, long-term care facility, or prison (21). The 2006 long-form census was a mandatory questionnaire with detailed questions pertaining to household demographic and socioeconomic information administered to 20% of Canadian households (21). In the 2011 census, the mandatory long-form questionnaires previously administered as part of the census were replaced with the NHS, a voluntary long-form questionnaire capturing the same demographic and socioeconomic information. The 2011 NHS was sent to 33% of households in Canada to account for expected lower response rates (22). Linkage to the DRD was achieved for 90.8% of the 2006 long-form census respondents and 96.7% of the 2011 NHS respondents (20).

#### Data sources

We linked the CanCHECs and their corresponding demographic data from the 2006 census and 2011 NHS to the Canadian Cancer Registry (CCR) using the DRD through 31 December 2015 (23), the Canadian Vital Statistics Death Registry (CVSD) through 31 December 2019 (24), and the T1 Personal Master File (T1PMF) through 31 December 2016 (25). The CCR is a national database containing all primary cancer diagnoses in Canada and corresponding tumour-specific information for individuals who reside in Canada, permanently or non-permanently (23). The CVSD is an administrative dataset that reports demographic and cause of death information for all deaths in Canada (24). The T1PMF is the annual postal code file containing mailing address postal codes for all Canadian residents who submitted a T1 income tax return form (20).

#### Cancer Diagnoses

To classify diagnoses into counts for each cancer site, we used the ICD-O-3 codes for SEER site groupings, except for head and neck cancers, for which we used the definitions from the 2021 Canadian Cancer Statistics (Supplementary Table 1) (26). Individuals could contribute multiple primary cancer diagnoses to analyses during follow-up. We used the International Agency for Research on Cancer rules for counting multiple primary cancers (27).

#### Socioeconomic and Demographic Variables

Because the CCR does not record demographic information other than age and sex, we used the 2006 long-form census and 2011 NHS for detailed, self-reported socioeconomic and demographic information, such as household after-tax income, household size, highest level of education, occupation industry sector, urbanicity, and race.

We used adjusted after-tax household income to derive individual-level income quintiles (28, 29). After-tax income data were collected for all individuals over 15 years of age living in private households. After-tax household income was calculated as the total income received by all household members occupying the same dwelling in the previous calendar year (2005 for the 2006 census and 2010 for the 2011 NHS) minus federal, provincial, and territorial income taxes paid in that year (28, 29). Non-taxable income was also included in total after-tax household income, except for income obtained via capital gains and losses, withdrawals from retirement savings plans and other savings plans intended to pay off debt, inheritances, lottery winnings, and insurance settlements (28, 29). For the 2006 long-form census and 2011 NHS, individuals could opt for permission to use income filed through their tax files, or, alternatively, to fill out the corresponding income-related questions in the questionnaires (28, 29). After-tax household income was adjusted by the square root of household size, which accounts for the increase in cost of material needs with increasing size (30). We used adjusted after-tax household income (28, 29) to rank individuals into individual-level income quintiles from lowest (Q1) to highest (Q5).

We used the Postal Code^OM^ Conversion File Plus, Version D (PCCF+) program to obtain area-level income quintiles based on individuals’ postal codes (31). The PCCF+ uses postal codes to probabilistically assign individuals to a census dissemination area (CDA). After assigning all respondents to their corresponding CDA, the median after-tax household income in each CDA was calculated based on 2006 census income data, multiplied by the number of households in the CDA, and divided by the single-person equivalents in the CDA to obtain the median after-tax income per single person equivalent. Based on the distribution of this measure in each CDA, the PCCF+ program derived area-level after-tax income quintiles for each census metropolitan area, census agglomeration, or residual area within each province. To account for individuals’ residential mobility and thus the time-varying nature of area-level income, we used annual postal codes from the T1PMF and assigned area-level income quintiles using the PCCF+ for each year an individual was alive from 2006 to 2015. In the years of an individual’s death or cancer diagnosis, we used the postal code from the CVSD or CCR respectively instead of the T1PMF, as these postal codes likely reflected more accurately an individual’s place of residence at the time of death or cancer diagnosis than their mailing address for tax filing purposes (25). Postal codes were not available for all cohort members or for all follow-up years due to late tax filings, missing tax records, and census respondents’ non-consent for tax return linkage (20). For years with missing postal code data, we imputed forwards the last known postal code or place of residence from the census. Income quintiles were designated as missing for individuals residing in some CDAs for which the PCCF+ could not assign area-level income due to missing area-level median income information (31).

### STATISTICAL ANALYSIS

The 2006 and 2011 CanCHECs were pooled to calculate age-standardized incidence rates (ASIR), age-standardized incidence rate ratios (ASIRR), and adjusted incidence rate ratios (IRR) using multivariable regression. We included person-time at risk and cancer diagnoses from the year of a person’s response to the questionnaire (i.e., 2006 for the census or 2011 for the NHS) until either their year of death or December 31, 2015. We excluded person-time contributed by Quebec residents after 2010 because new cancer diagnosis data from Quebec were not reported to the CCR after 2010. Analyses were conducted using SAS, version 9.4 (SAS Institute Inc).

#### Age-standardized rates and rate ratios

We calculated age-standardized incidence rates per 100,000 person-years and ASIRRs for cancers overall and by site using direct standardization to the 2011 Canadian census population as the standard population (32). We used the CanCHECs survey weights to calculate age-specific incidence rates representative of the Canadian population (20). The CanCHECs survey weights were created using existing census and NHS weights to account for differential non-linkage to the DRD as well as maintain the representativeness of the CanCHECs (20). We stratified age-standardized cancer incidence rates by individual- and area-level income quintile. We used the highest individual- and area-level income quintile (Q5) as the reference category for calculating ASIRRs. To estimate confidence intervals, we used the 500 bootstrap replicate weights developed for the 2006 and 2011 CanCHECs to account for sampling uncertainty (20). We used the 2.5 and 97.5 percentiles of the bootstrap distribution to generate 95% confidence intervals.

#### Multivariable negative binomial regression

We conducted multivariable negative binomial regression to model the independent associations between individual- and area-level income quintile, and cancer incidence by cancer site, adjusting for other demographic and socioeconomic variables. The models included the logarithm of the total person-years in a population subgroup as the offset. All models included both individual- and area-level income quintiles as independent variables, and were adjusted for 5-year age group, sex, year of observation, province of residence, urbanicity, visible minority group status, education level, and current occupation. Age, year of observation, province of residence, and area-level income quintile were treated as time-varying variables, to account for secular trends in cancer incidence and mobility, respectively. To calculate IRRs, we used the highest individual- and area-level income quintiles (Q5) as the reference categories; 95% confidence intervals were computed using the Wald method.

### ETHICS AND CONFIDENTIALITY

We obtained ethics approval from the McGill University Institutional Review Board for this analysis of secondary data. To protect respondent confidentiality, analysis outputs were vetted using rules developed by Statistics Canada, which include the rounding of all counts in tables and results to the nearest 5.

## RESULTS

Of the 2006 long-form census and 2011 NHS respondents, 5.9 million were included in the 2006 CanCHEC and 6.5 million were included in the 2011 CanCHEC (Table 1). In both cohorts, there were greater proportions of persons aged 14 years and below, as well as 65 years and above in the lowest individual-level household income quintile (Q1) compared to the middle (Q3) and highest (Q5) quintiles. Age distributions were stable across area-level income quintiles. The proportions of individuals who did not complete high school decreased, while the proportions of individuals who obtained a university or graduate degree increased with increasing income quintile. There were higher proportions of non-White and Indigenous peoples in the lowest individual- and area-level income quintiles compared to the highest. The proportions of individuals with rural residence decreased with increasing individual- and area-level income. The majority of individuals with missing area-level income data lived in rural areas (2006 CanCHEC, 72%; 2011 CanCHEC, 66%) rather than urban areas (2006 CanCHEC, 28%; CanCHEC 2011, 34%). The proportion of individuals employed in most occupational sectors generally tended to increase with individual- and area-level income quintile (Supplemental Table 2). This was likely because the variable for occupational sector was an indicator of current employment. Greater proportions of individuals who either were under 15 years of age, had not worked since the previous year, or had never worked were in the poorest individual- (2006 CanCHEC, 60%; 2011 CanCHEC, 61%) and area-level (2006 CanCHEC, 49%; 2011 CanCHEC, 49%) income quintiles compared to those in the wealthiest individual- (2006 CanCHEC, 24%; 2011 CanCHEC, 24%) and area-level (2006 CanCHEC, 38; 2011 CanCHEC, 39%) income quintiles, respectively.

**Table 1.**
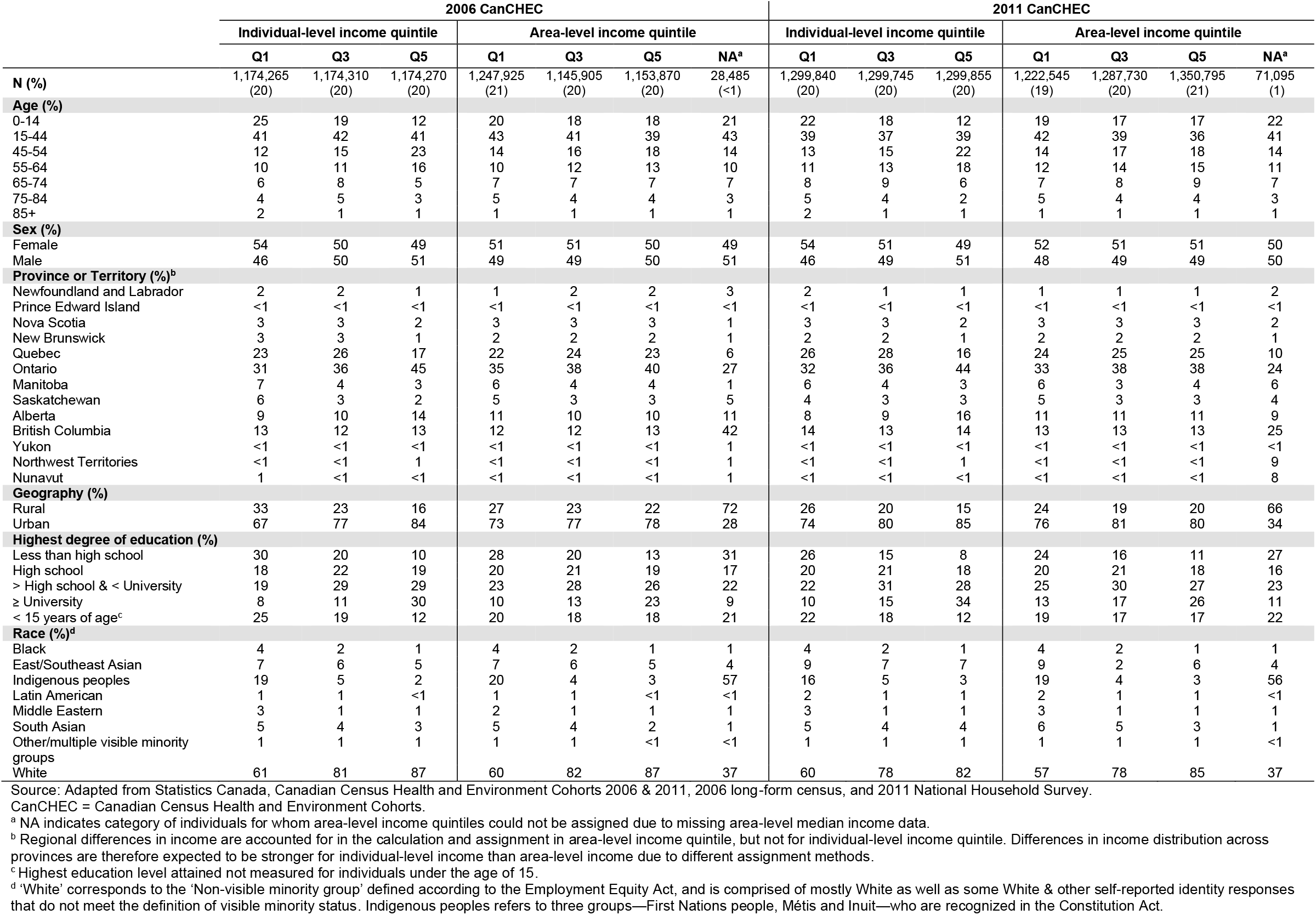
Baseline characteristics of 2006 and 2011 CanCHECs in lowest (Q1), middle (Q3), and highest (Q5) individual-level and area-level income quintiles

|  | 2006 CanCHEC |  |  |  |  |  |  | 2011 CanCHEC |  |  |  |  |  |  |
| --- | --- | --- | --- | --- | --- | --- | --- | --- | --- | --- | --- | --- | --- | --- |
|  | Individual-level income quintile |  |  | Area-level income quintile |  |  |  | Individual-level income quintile |  |  | Area-level income quintile |  |  |  |
|  | Q1 | Q3 | Q5 | Q1 | Q3 | Q5 | NA<sup>a</sup> | Q1 | Q3 | Q5 | Q1 | Q3 | Q5 | NA<sup>a</sup> |
| **N (%)** | 1,174,265<br>(20) | 1,174,310<br>(20) | 1,174,270<br>(20) | 1,247,925<br>(21) | 1,145,905<br>(20) | 1,153,870<br>(20) | 28,485<br>($<1$) | 1,299,840<br>(20) | 1,299,745<br>(20) | 1,299,855<br>(20) | 1,222,545<br>(19) | 1,287,730<br>(20) | 1,350,795<br>(21) | 71,095<br>(1) |
| **Age (%)** |  |  |  |  |  |  |  |  |  |  |  |  |  |  |
| 0-14 | 25 | 19 | 12 | 20 | 18 | 18 | 21 | 22 | 18 | 12 | 19 | 17 | 17 | 22 |
| 15-44 | 41 | 42 | 41 | 43 | 41 | 39 | 43 | 39 | 37 | 39 | 42 | 39 | 36 | 41 |
| 45-54 | 12 | 15 | 23 | 14 | 16 | 18 | 14 | 13 | 15 | 22 | 14 | 17 | 18 | 14 |
| 55-64 | 10 | 11 | 16 | 10 | 12 | 13 | 10 | 11 | 13 | 18 | 12 | 14 | 15 | 11 |
| 65-74 | 6 | 8 | 5 | 7 | 7 | 7 | 7 | 8 | 9 | 6 | 7 | 8 | 9 | 7 |
| 75-84 | 4 | 5 | 3 | 5 | 4 | 4 | 3 | 5 | 4 | 2 | 5 | 4 | 4 | 3 |
| 85+ | 2 | 1 | 1 | 1 | 1 | 1 | 1 | 2 | 1 | 1 | 1 | 1 | 1 | 1 |
| **Sex (%)** |  |  |  |  |  |  |  |  |  |  |  |  |  |  |
| Female | 54 | 50 | 49 | 51 | 51 | 50 | 49 | 54 | 51 | 49 | 52 | 51 | 51 | 50 |
| Male | 46 | 50 | 51 | 49 | 49 | 50 | 51 | 46 | 49 | 51 | 48 | 49 | 49 | 50 |
| **Province or Territory (%)<sup>b</sup>** |  |  |  |  |  |  |  |  |  |  |  |  |  |  |
| Newfoundland and Labrador | 2 | 2 | 1 | 1 | 2 | 2 | 3 | 2 | 1 | 1 | 1 | 1 | 1 | 2 |
| Prince Edward Island | $<1$ | $<1$ | $<1$ | $<1$ | $<1$ | $<1$ | $<1$ | $<1$ | $<1$ | $<1$ | $<1$ | $<1$ | $<1$ | $<1$ |
| Nova Scotia | 3 | 3 | 2 | 3 | 3 | 3 | 1 | 3 | 3 | 2 | 3 | 3 | 3 | 2 |
| New Brunswick | 3 | 3 | 1 | 2 | 2 | 2 | 1 | 2 | 2 | 1 | 2 | 2 | 2 | 1 |
| Quebec | 23 | 26 | 17 | 22 | 24 | 23 | 6 | 26 | 28 | 16 | 24 | 25 | 25 | 10 |
| Ontario | 31 | 36 | 45 | 35 | 38 | 40 | 27 | 32 | 36 | 44 | 33 | 38 | 38 | 24 |
| Manitoba | 7 | 4 | 3 | 6 | 4 | 4 | 1 | 6 | 4 | 3 | 6 | 3 | 4 | 6 |
| Saskatchewan | 6 | 3 | 2 | 5 | 3 | 3 | 5 | 4 | 3 | 3 | 5 | 3 | 3 | 4 |
| Alberta | 9 | 10 | 14 | 11 | 10 | 10 | 11 | 8 | 9 | 16 | 11 | 11 | 11 | 9 |
| British Columbia | 13 | 12 | 13 | 12 | 12 | 13 | 42 | 14 | 13 | 14 | 13 | 13 | 13 | 25 |
| Yukon | $<1$ | $<1$ | $<1$ | $<1$ | $<1$ | $<1$ | 1 | $<1$ | $<1$ | $<1$ | $<1$ | $<1$ | $<1$ | $<1$ |
| Northwest Territories | $<1$ | $<1$ | 1 | $<1$ | $<1$ | $<1$ | 1 | $<1$ | $<1$ | 1 | $<1$ | $<1$ | $<1$ | 9 |
| Nunavut | 1 | $<1$ | $<1$ | $<1$ | $<1$ | $<1$ | 1 | $<1$ | $<1$ | $<1$ | $<1$ | $<1$ | $<1$ | 8 |
| **Geography (%)** |  |  |  |  |  |  |  |  |  |  |  |  |  |  |
| Rural | 33 | 23 | 16 | 27 | 23 | 22 | 72 | 26 | 20 | 15 | 24 | 19 | 20 | 66 |
| Urban | 67 | 77 | 84 | 73 | 77 | 78 | 28 | 74 | 80 | 85 | 76 | 81 | 80 | 34 |
| **Highest degree of education (%)** |  |  |  |  |  |  |  |  |  |  |  |  |  |  |
| Less than high school | 30 | 20 | 10 | 28 | 20 | 13 | 31 | 26 | 15 | 8 | 24 | 16 | 11 | 27 |
| High school | 18 | 22 | 19 | 20 | 21 | 19 | 17 | 20 | 21 | 18 | 20 | 21 | 18 | 16 |
| > High school & < University | 19 | 29 | 29 | 23 | 28 | 26 | 22 | 22 | 31 | 28 | 25 | 30 | 27 | 23 |
| ≥ University | 8 | 11 | 30 | 10 | 13 | 23 | 9 | 10 | 15 | 34 | 13 | 17 | 26 | 11 |
| < 15 years of age<sup>c</sup> | 25 | 19 | 12 | 20 | 18 | 18 | 21 | 22 | 18 | 12 | 19 | 17 | 17 | 22 |
| **Race (%)<sup>d</sup>** |  |  |  |  |  |  |  |  |  |  |  |  |  |  |
| Black | 4 | 2 | 1 | 4 | 2 | 1 | 1 | 4 | 2 | 1 | 4 | 2 | 1 | 1 |
| East/Southeast Asian | 7 | 6 | 5 | 7 | 6 | 5 | 4 | 9 | 7 | 7 | 9 | 2 | 6 | 4 |
| Indigenous peoples | 19 | 5 | 2 | 20 | 4 | 3 | 57 | 16 | 5 | 3 | 19 | 4 | 3 | 56 |
| Latin American | 1 | 1 | $<1$ | 1 | 1 | $<1$ | $<1$ | 2 | 1 | 1 | 2 | 1 | 1 | $<1$ |
| Middle Eastern | 3 | 1 | 1 | 2 | 1 | 1 | 1 | 3 | 1 | 1 | 3 | 1 | 1 | 1 |
| South Asian | 5 | 4 | 3 | 5 | 4 | 2 | 1 | 5 | 4 | 4 | 6 | 5 | 3 | 1 |
| Other/multiple visible minority groups | 1 | 1 | 1 | 1 | 1 | $<1$ | $<1$ | 1 | 1 | 1 | 1 | 1 | 1 | $<1$ |
| White | 61 | 81 | 87 | 60 | 82 | 87 | 37 | 60 | 78 | 82 | 57 | 78 | 85 | 37 |

Source: Adapted from Statistics Canada, Canadian Census Health and Environment Cohorts 2006 & 2011, 2006 long-form census, and 2011 National Household Survey.
CanCHEC = Canadian Census Health and Environment Cohorts.
<sup>a</sup> NA indicates category of individuals for whom area-level income quintiles could not be assigned due to missing area-level median income data.<sup>b</sup> Regional differences in income are accounted for in the calculation and assignment in area-level income quintile, but not for individual-level income quintile. Differences in income distribution across provinces are therefore expected to be stronger for individual-level income than area-level income due to different assignment methods.<sup>c</sup> Highest education level attained not measured for individuals under the age of 15.<sup>d</sup> 'White' corresponds to the 'Non-visible minority group' defined according to the Employment Equity Act, and is comprised of mostly White as well as some White & other self-reported identity responses that do not meet the definition of visible minority status. Indigenous peoples refers to three groups—First Nations people, Métis and Inuit—who are recognized in the Constitution Act.

Age-standardized overall cancer incidence rates and ASIRRs by individual- and area-level income quintile are presented in Table 2. Cumulatively, 411,220 cancers were diagnosed during follow-up. Overall age standardized cancer incidence rates decreased as individual- and area-level income increased. Individuals who were both in the combined poorest individual- and area-level income quintiles had significantly higher overall age-standardized cancer incidence rates compared to those in the combined wealthiest individual- and area-level income quintiles (ASIRR 1.09, 95% CI 1.07-1.12).

**Table 2.** Age-standardized overall cancer incidence rates per 100,000 person-years, age-standardized incidence rate ratios, and 95% CIs by individual- and area-level income quintile, 2006 and 2011 CanCHECs combined and weighted, standardized to the Canadian 2011 census population

|  | Age-Standardized Incidence Rates (/100,000 person-years) |  |  |  |  |  | Age-Standardized Incidence Rate Ratios |  |  |  |  |
| --- | --- | --- | --- | --- | --- | --- | --- | --- | --- | --- | --- |
|  | Individual-level income quintile |  |  |  |  |  | Individual-level income quintile |  |  |  |  |
| Area-level income quintile | Q1 | Q2 | Q3 | Q4 | Q5 | Overall | Q1 | Q2 | Q3 | Q4 | Q5 |
| **Q1** | 542 | 533 | 537 | 525 | 512 | 534 | **1.09** | **1.07** | **1.08** | **1.06** | **1.03** |
| 95% CI | (534-552) | (524-541) | (525-547) | (513-538) | (495-528) | (529-539) | **(1.07-1.12)** | **(1.05-1.09)** | **(1.05-1.10)** | **(1.03-1.09)** | **(1.00-1.07)** |
| **Q2** | 520 | 527 | 518 | 519 | 511 | 520 | **1.05** | **1.06** | **1.04** | **1.04** | **1.03** |
| 95% CI | (510-532) | (518-535) | (509-527) | (510-529) | (499-523) | (516-524) | **(1.02-1.07)** | **(1.04-1.06)** | **(1.02-1.07)** | **(1.02-1.07)** | **(1.00-1.06)** |
| **Q3** | 507 | 516 | 519 | 511 | 512 | 513 | 1.02 | **1.04** | **1.04** | **1.03** | **1.03** |
| 95% CI | (495-517) | (505-524) | (510-529) | (501-520) | (502-521) | (509-517) | (0.99-1.04) | **(1.01-1.06)** | **(1.02-1.07)** | **(1.00-1.05)** | **(1.01-1.05)** |
| **Q4** | 490 | 520 | 508 | 506 | 500 | 505 | 0.99 | **1.05** | **1.02** | 1.02 | 1.01 |
| 95% CI | (477-502) | (509-530) | (499-518) | (497-515) | (492-508) | (501-509) | (0.96-1.02) | **(1.02-1.07)** | **(1.00-1.05)** | (0.99-1.04) | (0.99-1.03) |
| **Q5** | 497 | 515 | 511 | 499 | 497 | 503 | 1.00 | **1.04** | **1.03** | 1.00 | 1.00 |
| 95% CI | (483-512) | (503-525) | (502-522) | (490-508) | (491-504) | (499-507) | (0.97-1.03) | **(1.01-1.06)** | **(1.00-1.06)** | (0.98-1.02) | (Ref) |
| **Overall** | 519 | 523 | 518 | 510 | 503 | 514 |  |  |  |  |  |
| 95% CI | (513-523) | (519-527) | (514-522) | (506-515) | (499-507) | (512-516) |  |  |  |  |  |

Source: Adapted from Statistics Canada, Canadian Census Health and Environment Cohorts 2006 & 2011, Canadian Cancer Registry, 2006 long-form census, and 2011 National Household Survey.
95% CI = confidence intervals based on 2.5<sup>th</sup> and 97.5<sup>th</sup> bootstrap percentiles.
Bolded age-standardized incidence rate ratio estimates are significant based on 95% confidence intervals.

Table 3 and Table 4 present site-specific ASIRs and ASIRRs by individual- and area-level income quintiles, respectively. Individuals in the lowest individual- and area-level income quintiles had higher age-standardized incidence rates compared to those in the highest income quintiles for head and neck, oropharyngeal, esophageal, stomach, colorectal, anal, liver, pancreas, lung, cervical, and kidney and renal pelvis cancers (ASIRRs for lowest versus highest quintiles ranged from 1.01 to 1.92). Conversely, age-standardized incidence rates increased as individual- and area-level income increased for breast, uterine, prostate, and testicular cancers, as well as for melanomas, Hodgkin’s lymphomas (HL), and non-Hodgkin’s lymphomas (NHL) (ASIRRs for lowest versus highest quintiles ranged from 0.45 to 0.97). The age-standardized bladder cancer incidence rate was lower for individuals in the poorest individual-level income quintile compared to those in the wealthiest individual level-income quintile (ASIRR 0.92, 95% CI 0.87-0.98), but higher in the poorest area-level income quintile than in the wealthiest area-level income quintile (ASIRR 1.05, 95% CI 1.00-1.11). Individuals in the poorest individual-level income quintile had higher age-standardized incidence rates of multiple myeloma and leukemia than individuals in the wealthiest individual-level income quintile, while age-standardized incidence rates for these cancers were comparable across area-level income quintiles. For thyroid as well as brain and central nervous system (CNS) cancers, the age standardized incidence rates were lower in the poorest area-level income quintile compared to those in the wealthiest area-level income quintile, but incidence rates were comparable across individual-level income quintiles.

**Table 3.**
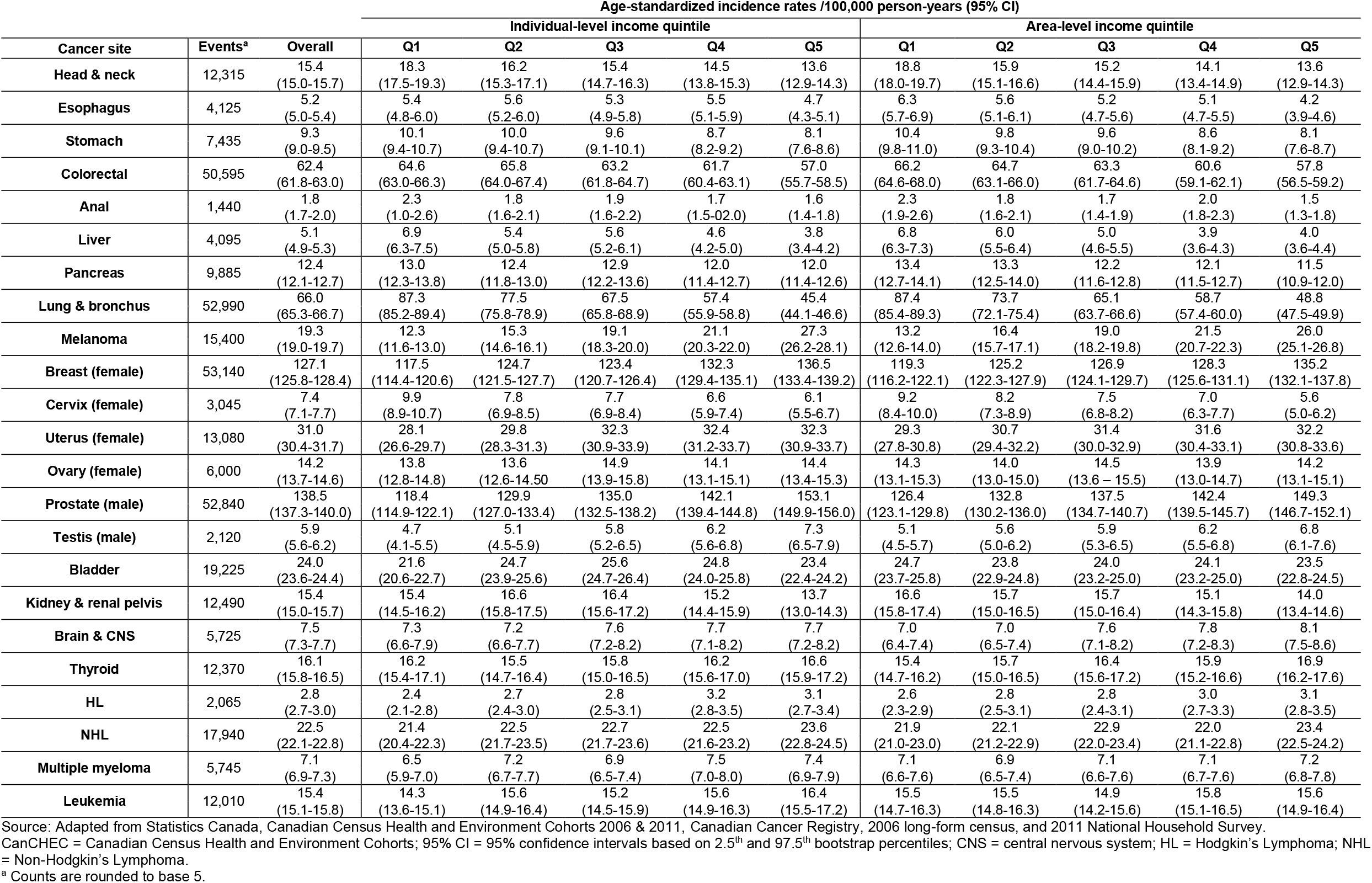
Age-standardized site-specific cancer incidence rates per 100,000 person-years and 95% CIs by individual- and area-level income quintile, 2006 and 2011 CanCHECs combined and weighted, standardized to the Canadian 2011 census population

**Table 4.**
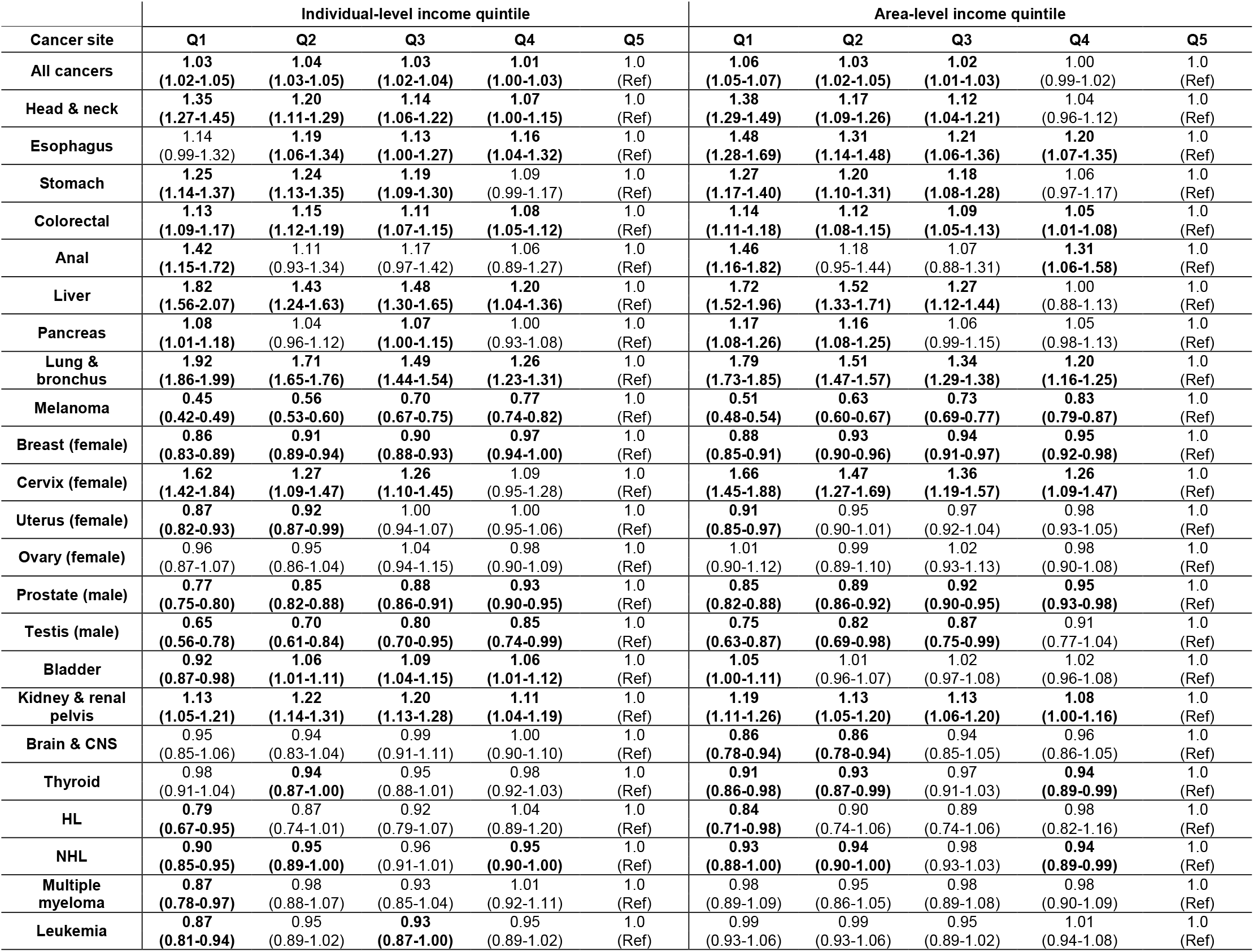
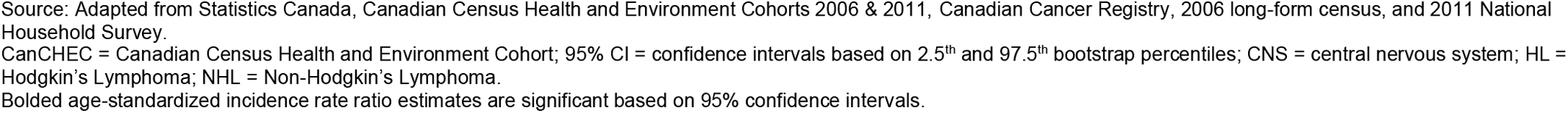
Age-standardized site-specific incidence rate ratios and 95% CIs by individual- and area-level income quintile, 2006 and 2011 CanCHECs combined and weighted, standardized to the Canadian 2011 census population

| Cancer site | Individual-level income quintile |  |  |  |  | Area-level income quintile |  |  |  |  |
| --- | --- | --- | --- | --- | --- | --- | --- | --- | --- | --- |
|  | Q1 | Q2 | Q3 | Q4 | Q5 | Q1 | Q2 | Q3 | Q4 | Q5 |
| All cancers | 1.03<br>(1.02-1.05) | 1.04<br>(1.03-1.05) | 1.03<br>(1.02-1.04) | 1.01<br>(1.00-1.03) | 1.0<br>(Ref) | 1.06<br>(1.05-1.07) | 1.03<br>(1.02-1.05) | 1.02<br>(1.01-1.03) | 1.00<br>(0.99-1.02) | 1.0<br>(Ref) |
| Head & neck | 1.35<br>(1.27-1.45) | 1.20<br>(1.11-1.29) | 1.14<br>(1.06-1.22) | 1.07<br>(1.00-1.15) | 1.0<br>(Ref) | 1.38<br>(1.29-1.49) | 1.17<br>(1.09-1.26) | 1.12<br>(1.04-1.21) | 1.04<br>(0.96-1.12) | 1.0<br>(Ref) |
| Esophagus | 1.14<br>(0.99-1.32) | 1.19<br>(1.06-1.34) | 1.13<br>(1.00-1.27) | 1.16<br>(1.04-1.32) | 1.0<br>(Ref) | 1.48<br>(1.28-1.69) | 1.31<br>(1.14-1.48) | 1.21<br>(1.06-1.36) | 1.20<br>(1.07-1.35) | 1.0<br>(Ref) |
| Stomach | 1.25<br>(1.14-1.37) | 1.24<br>(1.13-1.35) | 1.19<br>(1.09-1.30) | 1.09<br>(0.99-1.17) | 1.0<br>(Ref) | 1.27<br>(1.17-1.40) | 1.20<br>(1.10-1.31) | 1.18<br>(1.08-1.28) | 1.06<br>(0.97-1.17) | 1.0<br>(Ref) |
| Colorectal | 1.13<br>(1.09-1.17) | 1.15<br>(1.12-1.19) | 1.11<br>(1.07-1.15) | 1.08<br>(1.05-1.12) | 1.0<br>(Ref) | 1.14<br>(1.11-1.18) | 1.12<br>(1.08-1.15) | 1.09<br>(1.05-1.13) | 1.05<br>(1.01-1.08) | 1.0<br>(Ref) |
| Anal | 1.42<br>(1.15-1.72) | 1.11<br>(0.93-1.34) | 1.17<br>(0.97-1.42) | 1.06<br>(0.89-1.27) | 1.0<br>(Ref) | 1.46<br>(1.16-1.82) | 1.18<br>(0.95-1.44) | 1.07<br>(0.88-1.31) | 1.31<br>(1.06-1.58) | 1.0<br>(Ref) |
| Liver | 1.82<br>(1.56-2.07) | 1.43<br>(1.24-1.63) | 1.48<br>(1.30-1.65) | 1.20<br>(1.04-1.36) | 1.0<br>(Ref) | 1.72<br>(1.52-1.96) | 1.52<br>(1.33-1.71) | 1.27<br>(1.12-1.44) | 1.00<br>(0.88-1.13) | 1.0<br>(Ref) |
| Pancreas | 1.08<br>(1.01-1.18) | 1.04<br>(0.96-1.12) | 1.07<br>(1.00-1.15) | 1.00<br>(0.93-1.08) | 1.0<br>(Ref) | 1.17<br>(1.08-1.26) | 1.16<br>(1.08-1.25) | 1.06<br>(0.99-1.15) | 1.05<br>(0.98-1.13) | 1.0<br>(Ref) |
| Lung & bronchus | 1.92<br>(1.86-1.99) | 1.71<br>(1.65-1.76) | 1.49<br>(1.44-1.54) | 1.26<br>(1.23-1.31) | 1.0<br>(Ref) | 1.79<br>(1.73-1.85) | 1.51<br>(1.47-1.57) | 1.34<br>(1.29-1.38) | 1.20<br>(1.16-1.25) | 1.0<br>(Ref) |
| Melanoma | 0.45<br>(0.42-0.49) | 0.56<br>(0.53-0.60) | 0.70<br>(0.67-0.75) | 0.77<br>(0.74-0.82) | 1.0<br>(Ref) | 0.51<br>(0.48-0.54) | 0.63<br>(0.60-0.67) | 0.73<br>(0.69-0.77) | 0.83<br>(0.79-0.87) | 1.0<br>(Ref) |
| Breast (female) | 0.86<br>(0.83-0.89) | 0.91<br>(0.89-0.94) | 0.90<br>(0.88-0.93) | 0.97<br>(0.94-1.00) | 1.0<br>(Ref) | 0.88<br>(0.85-0.91) | 0.93<br>(0.90-0.96) | 0.94<br>(0.91-0.97) | 0.95<br>(0.92-0.98) | 1.0<br>(Ref) |
| Cervix (female) | 1.62<br>(1.42-1.84) | 1.27<br>(1.09-1.47) | 1.26<br>(1.10-1.45) | 1.09<br>(0.95-1.28) | 1.0<br>(Ref) | 1.66<br>(1.45-1.88) | 1.47<br>(1.27-1.69) | 1.36<br>(1.19-1.57) | 1.26<br>(1.09-1.47) | 1.0<br>(Ref) |
| Uterus (female) | 0.87<br>(0.82-0.93) | 0.92<br>(0.87-0.99) | 1.00<br>(0.94-1.07) | 1.00<br>(0.95-1.06) | 1.0<br>(Ref) | 0.91<br>(0.85-0.97) | 0.95<br>(0.90-1.01) | 0.97<br>(0.92-1.04) | 0.98<br>(0.93-1.05) | 1.0<br>(Ref) |
| Ovary (female) | 0.96<br>(0.87-1.07) | 0.95<br>(0.86-1.04) | 1.04<br>(0.94-1.15) | 0.98<br>(0.90-1.09) | 1.0<br>(Ref) | 1.01<br>(0.90-1.12) | 0.99<br>(0.89-1.10) | 1.02<br>(0.93-1.13) | 0.98<br>(0.90-1.08) | 1.0<br>(Ref) |
| Prostate (male) | 0.77<br>(0.75-0.80) | 0.85<br>(0.82-0.88) | 0.88<br>(0.86-0.91) | 0.93<br>(0.90-0.95) | 1.0<br>(Ref) | 0.85<br>(0.82-0.88) | 0.89<br>(0.86-0.92) | 0.92<br>(0.90-0.95) | 0.95<br>(0.93-0.98) | 1.0<br>(Ref) |
| Testis (male) | 0.65<br>(0.56-0.78) | 0.70<br>(0.61-0.84) | 0.80<br>(0.70-0.95) | 0.85<br>(0.74-0.99) | 1.0<br>(Ref) | 0.75<br>(0.63-0.87) | 0.82<br>(0.69-0.98) | 0.87<br>(0.75-0.99) | 0.91<br>(0.77-1.04) | 1.0<br>(Ref) |
| Bladder | 0.92<br>(0.87-0.98) | 1.06<br>(1.01-1.11) | 1.09<br>(1.04-1.15) | 1.06<br>(1.01-1.12) | 1.0<br>(Ref) | 1.05<br>(1.00-1.11) | 1.01<br>(0.96-1.07) | 1.02<br>(0.97-1.08) | 1.02<br>(0.96-1.08) | 1.0<br>(Ref) |
| Kidney & renal pelvis | 1.13<br>(1.05-1.21) | 1.22<br>(1.14-1.31) | 1.20<br>(1.13-1.28) | 1.11<br>(1.04-1.19) | 1.0<br>(Ref) | 1.19<br>(1.11-1.26) | 1.13<br>(1.05-1.20) | 1.13<br>(1.06-1.20) | 1.08<br>(1.00-1.16) | 1.0<br>(Ref) |
| Brain & CNS | 0.95<br>(0.85-1.06) | 0.94<br>(0.83-1.04) | 0.99<br>(0.91-1.11) | 1.00<br>(0.90-1.10) | 1.0<br>(Ref) | 0.86<br>(0.78-0.94) | 0.86<br>(0.78-0.94) | 0.94<br>(0.85-1.05) | 0.96<br>(0.86-1.05) | 1.0<br>(Ref) |
| Thyroid | 0.98<br>(0.91-1.04) | 0.94<br>(0.87-1.00) | 0.95<br>(0.88-1.01) | 0.98<br>(0.92-1.03) | 1.0<br>(Ref) | 0.91<br>(0.86-0.98) | 0.93<br>(0.87-0.99) | 0.97<br>(0.91-1.03) | 0.94<br>(0.89-0.99) | 1.0<br>(Ref) |
| HL | 0.79<br>(0.67-0.95) | 0.87<br>(0.74-1.01) | 0.92<br>(0.79-1.07) | 1.04<br>(0.89-1.20) | 1.0<br>(Ref) | 0.84<br>(0.71-0.98) | 0.90<br>(0.74-1.06) | 0.89<br>(0.74-1.06) | 0.98<br>(0.82-1.16) | 1.0<br>(Ref) |
| NHL | 0.90<br>(0.85-0.95) | 0.95<br>(0.89-1.00) | 0.96<br>(0.91-1.01) | 0.95<br>(0.90-1.00) | 1.0<br>(Ref) | 0.93<br>(0.88-1.00) | 0.94<br>(0.90-1.00) | 0.98<br>(0.93-1.03) | 0.94<br>(0.89-0.99) | 1.0<br>(Ref) |
| Multiple myeloma | 0.87<br>(0.78-0.97) | 0.98<br>(0.88-1.07) | 0.93<br>(0.85-1.04) | 1.01<br>(0.92-1.11) | 1.0<br>(Ref) | 0.98<br>(0.89-1.09) | 0.95<br>(0.86-1.05) | 0.98<br>(0.89-1.08) | 0.98<br>(0.90-1.09) | 1.0<br>(Ref) |
| Leukemia | 0.87<br>(0.81-0.94) | 0.95<br>(0.89-1.02) | 0.93<br>(0.87-1.00) | 0.95<br>(0.89-1.02) | 1.0<br>(Ref) | 0.99<br>(0.93-1.06) | 0.99<br>(0.93-1.06) | 0.95<br>(0.89-1.02) | 1.01<br>(0.94-1.08) | 1.0<br>(Ref) |

Source: Adapted from Statistics Canada, Canadian Census Health and Environment Cohorts 2006 & 2011, Canadian Cancer Registry, 2006 long-form census, and 2011 National Household Survey.
CanCHEC = Canadian Census Health and Environment Cohort; 95% CI = confidence intervals based on 2.5<sup>th</sup> and 97.5<sup>th</sup> bootstrap percentiles; CNS = central nervous system; HL = Hodgkin's Lymphoma; NHL = Non-Hodgkin's Lymphoma.
Bolded age-standardized incidence rate ratio estimates are significant based on 95% confidence intervals.

After mutually adjusting incidence rates for both individual- and area-level income as well as for other elements of SES, the independent associations between individual- and area-level income, and site-specific cancer incidence were slightly attenuated but remained significant for most cancer sites (Table 5). Adjusted incidence rates persisted in being higher in poorer individual- and area-level income quintiles for head and neck, esophageal, stomach, colorectal, anal, liver, lung, cervical, and kidney and renal pelvis cancers. Inversely, adjusted incidence rates stayed lower among poorer individual- and area-level income quintiles for melanoma, breast, prostate, testicular, and thyroid cancers. The associations of income with HL, NHL, and leukemia incidence, however, largely disappeared after adjustment. For brain and CNS cancers, incidence remained lower for individuals in poorer area-level income quintiles following adjustment, whereas individual-level income continued to have no association. After adjustment, incidence rates of pancreatic cancer among individuals in lower area-level income quintiles remained higher, but the effect of individual-level income was attenuated and no longer significant. The previously observed decreased age-standardized uterine cancer incidence among lower individual- and area-level income quintiles increased for middle individual-level income quintiles and association between of area-level income and uterine cancer incidence was eliminated with adjustment. Adjustment revealed higher bladder cancer incidence among lower individual-level income quintiles, which had been lower prior to adjustment, and also increased the magnitude of higher bladder cancer incidence among lower area-level income quintiles. Adjusted ovarian cancer incidence increased in magnitude for area-level income quintiles, while adjusted ovarian cancer incidence remained equivalent across the individual-level income gradient.

**Table 5.**
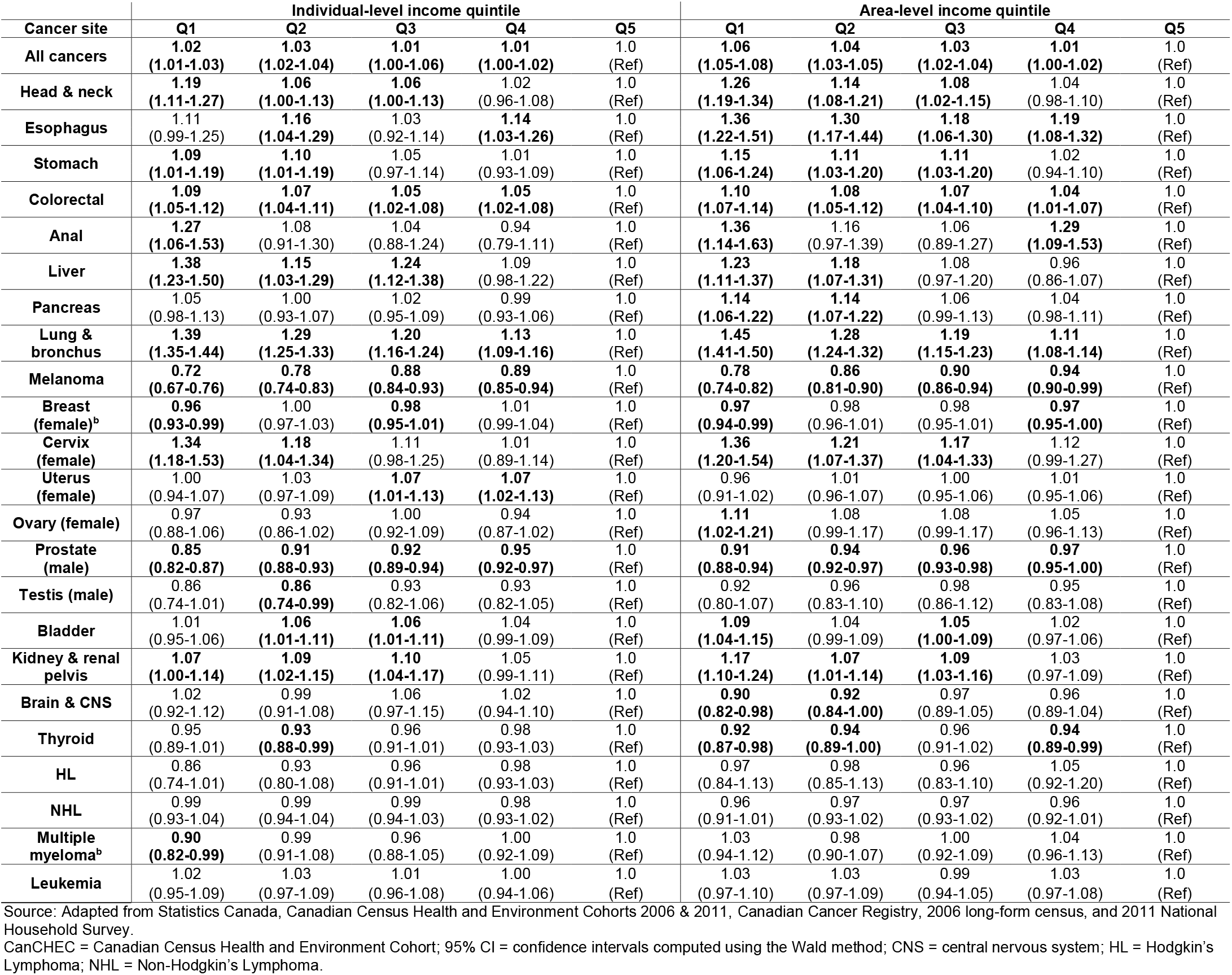
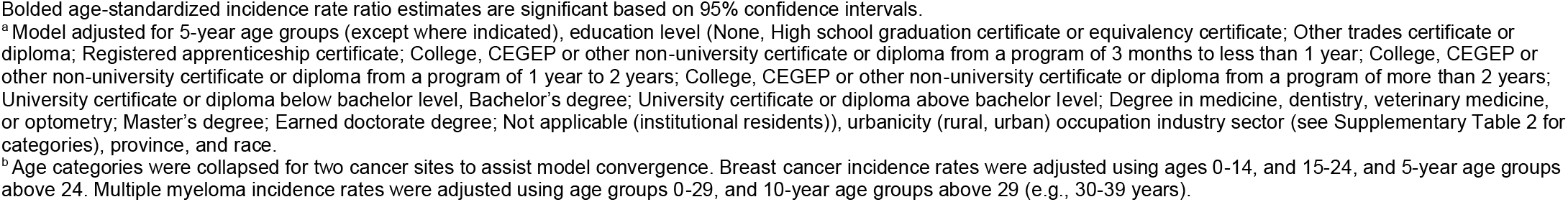
Overall and site-specific adjusteda multivariable negative binomial regression cancer incidence rate ratios (95% CIs) by individual- and area-level income quintile

| Cancer site | Individual-level income quintile |  |  |  |  | Area-level income quintile |  |  |  |  |
| --- | --- | --- | --- | --- | --- | --- | --- | --- | --- | --- |
|  | Q1 | Q2 | Q3 | Q4 | Q5 | Q1 | Q2 | Q3 | Q4 | Q5 |
| All cancers | 1.02<br>(1.01-1.03) | 1.03<br>(1.02-1.04) | 1.01<br>(1.00-1.06) | 1.01<br>(1.00-1.02) | 1.0<br>(Ref) | 1.06<br>(1.05-1.08) | 1.04<br>(1.03-1.05) | 1.03<br>(1.02-1.04) | 1.01<br>(1.00-1.02) | 1.0<br>(Ref) |
| Head & neck | 1.19<br>(1.11-1.27) | 1.06<br>(1.00-1.13) | 1.06<br>(1.00-1.13) | 1.02<br>(0.96-1.08) | 1.0<br>(Ref) | 1.26<br>(1.19-1.34) | 1.14<br>(1.08-1.21) | 1.08<br>(1.02-1.15) | 1.04<br>(0.98-1.10) | 1.0<br>(Ref) |
| Esophagus | 1.11<br>(0.99-1.25) | 1.16<br>(1.04-1.29) | 1.03<br>(0.92-1.14) | 1.14<br>(1.03-1.26) | 1.0<br>(Ref) | 1.36<br>(1.22-1.51) | 1.30<br>(1.17-1.44) | 1.18<br>(1.06-1.30) | 1.19<br>(1.08-1.32) | 1.0<br>(Ref) |
| Stomach | 1.09<br>(1.01-1.19) | 1.10<br>(1.01-1.19) | 1.05<br>(0.97-1.14) | 1.01<br>(0.93-1.09) | 1.0<br>(Ref) | 1.15<br>(1.06-1.24) | 1.11<br>(1.03-1.20) | 1.11<br>(1.03-1.20) | 1.02<br>(0.94-1.10) | 1.0<br>(Ref) |
| Colorectal | 1.09<br>(1.05-1.12) | 1.07<br>(1.04-1.11) | 1.05<br>(1.02-1.08) | 1.05<br>(1.02-1.08) | 1.0<br>(Ref) | 1.10<br>(1.07-1.14) | 1.08<br>(1.05-1.12) | 1.07<br>(1.04-1.10) | 1.04<br>(1.01-1.07) | 1.0<br>(Ref) |
| Anal | 1.27<br>(1.06-1.53) | 1.08<br>(0.91-1.30) | 1.04<br>(0.88-1.24) | 0.94<br>(0.79-1.11) | 1.0<br>(Ref) | 1.36<br>(1.14-1.63) | 1.16<br>(0.97-1.39) | 1.06<br>(0.89-1.27) | 1.29<br>(1.09-1.53) | 1.0<br>(Ref) |
| Liver | 1.38<br>(1.23-1.50) | 1.15<br>(1.03-1.29) | 1.24<br>(1.12-1.38) | 1.09<br>(0.98-1.22) | 1.0<br>(Ref) | 1.23<br>(1.11-1.37) | 1.18<br>(1.07-1.31) | 1.08<br>(0.97-1.20) | 0.96<br>(0.86-1.07) | 1.0<br>(Ref) |
| Pancreas | 1.05<br>(0.98-1.13) | 1.00<br>(0.93-1.07) | 1.02<br>(0.95-1.09) | 0.99<br>(0.93-1.06) | 1.0<br>(Ref) | 1.14<br>(1.06-1.22) | 1.14<br>(1.07-1.22) | 1.06<br>(0.99-1.13) | 1.04<br>(0.98-1.11) | 1.0<br>(Ref) |
| Lung & bronchus | 1.39<br>(1.35-1.44) | 1.29<br>(1.25-1.33) | 1.20<br>(1.16-1.24) | 1.13<br>(1.09-1.16) | 1.0<br>(Ref) | 1.45<br>(1.41-1.50) | 1.28<br>(1.24-1.32) | 1.19<br>(1.15-1.23) | 1.11<br>(1.08-1.14) | 1.0<br>(Ref) |
| Melanoma | 0.72<br>(0.67-0.76) | 0.78<br>(0.74-0.83) | 0.88<br>(0.84-0.93) | 0.89<br>(0.85-0.94) | 1.0<br>(Ref) | 0.78<br>(0.74-0.82) | 0.86<br>(0.81-0.90) | 0.90<br>(0.86-0.94) | 0.94<br>(0.90-0.99) | 1.0<br>(Ref) |
| Breast (female)<sup>b</sup> | 0.96<br>(0.93-0.99) | 1.00<br>(0.97-1.03) | 0.98<br>(0.95-1.01) | 1.01<br>(0.99-1.04) | 1.0<br>(Ref) | 0.97<br>(0.94-0.99) | 0.98<br>(0.96-1.01) | 0.98<br>(0.95-1.01) | 0.97<br>(0.95-1.00) | 1.0<br>(Ref) |
| Cervix (female) | 1.34<br>(1.18-1.53) | 1.18<br>(1.04-1.34) | 1.11<br>(0.98-1.25) | 1.01<br>(0.89-1.14) | 1.0<br>(Ref) | 1.36<br>(1.20-1.54) | 1.21<br>(1.07-1.37) | 1.17<br>(1.04-1.33) | 1.12<br>(0.99-1.27) | 1.0<br>(Ref) |
| Uterus (female) | 1.00<br>(0.94-1.07) | 1.03<br>(0.97-1.09) | 1.07<br>(1.01-1.13) | 1.07<br>(1.02-1.13) | 1.0<br>(Ref) | 0.96<br>(0.91-1.02) | 1.01<br>(0.96-1.07) | 1.00<br>(0.95-1.06) | 1.01<br>(0.95-1.06) | 1.0<br>(Ref) |
| Ovary (female) | 0.97<br>(0.88-1.06) | 0.93<br>(0.86-1.02) | 1.00<br>(0.92-1.09) | 0.94<br>(0.87-1.02) | 1.0<br>(Ref) | 1.11<br>(1.02-1.21) | 1.08<br>(0.99-1.17) | 1.08<br>(0.99-1.17) | 1.05<br>(0.96-1.13) | 1.0<br>(Ref) |
| Prostate (male) | 0.85<br>(0.82-0.87) | 0.91<br>(0.88-0.93) | 0.92<br>(0.89-0.94) | 0.95<br>(0.92-0.97) | 1.0<br>(Ref) | 0.91<br>(0.88-0.94) | 0.94<br>(0.92-0.97) | 0.96<br>(0.93-0.98) | 0.97<br>(0.95-1.00) | 1.0<br>(Ref) |
| Testis (male) | 0.86<br>(0.74-1.01) | 0.86<br>(0.74-0.99) | 0.93<br>(0.82-1.06) | 0.93<br>(0.82-1.05) | 1.0<br>(Ref) | 0.92<br>(0.80-1.07) | 0.96<br>(0.83-1.10) | 0.98<br>(0.86-1.12) | 0.95<br>(0.83-1.08) | 1.0<br>(Ref) |
| Bladder | 1.01<br>(0.95-1.06) | 1.06<br>(1.01-1.11) | 1.06<br>(1.01-1.11) | 1.04<br>(0.99-1.09) | 1.0<br>(Ref) | 1.09<br>(1.04-1.15) | 1.04<br>(0.99-1.09) | 1.05<br>(1.00-1.09) | 1.02<br>(0.97-1.06) | 1.0<br>(Ref) |
| Kidney & renal pelvis | 1.07<br>(1.00-1.14) | 1.09<br>(1.02-1.15) | 1.10<br>(1.04-1.17) | 1.05<br>(0.99-1.11) | 1.0<br>(Ref) | 1.17<br>(1.10-1.24) | 1.07<br>(1.01-1.14) | 1.09<br>(1.03-1.16) | 1.03<br>(0.97-1.09) | 1.0<br>(Ref) |
| Brain & CNS | 1.02<br>(0.92-1.12) | 0.99<br>(0.91-1.08) | 1.06<br>(0.97-1.15) | 1.02<br>(0.94-1.10) | 1.0<br>(Ref) | 0.90<br>(0.82-0.98) | 0.92<br>(0.84-1.00) | 0.97<br>(0.89-1.05) | 0.96<br>(0.89-1.04) | 1.0<br>(Ref) |
| Thyroid | 0.95<br>(0.89-1.01) | 0.93<br>(0.88-0.99) | 0.96<br>(0.91-1.01) | 0.98<br>(0.93-1.03) | 1.0<br>(Ref) | 0.92<br>(0.87-0.98) | 0.94<br>(0.89-1.00) | 0.96<br>(0.91-1.02) | 0.94<br>(0.89-0.99) | 1.0<br>(Ref) |
| HL | 0.86<br>(0.74-1.01) | 0.93<br>(0.80-1.08) | 0.96<br>(0.91-1.01) | 0.98<br>(0.93-1.03) | 1.0<br>(Ref) | 0.97<br>(0.84-1.13) | 0.98<br>(0.85-1.13) | 0.96<br>(0.83-1.10) | 1.05<br>(0.92-1.20) | 1.0<br>(Ref) |
| NHL | 0.99<br>(0.93-1.04) | 0.99<br>(0.94-1.04) | 0.99<br>(0.94-1.03) | 0.98<br>(0.93-1.02) | 1.0<br>(Ref) | 0.96<br>(0.91-1.01) | 0.97<br>(0.93-1.02) | 0.97<br>(0.93-1.02) | 0.96<br>(0.92-1.01) | 1.0<br>(Ref) |
| Multiple myeloma<sup>b</sup> | 0.90<br>(0.82-0.99) | 0.99<br>(0.91-1.08) | 0.96<br>(0.88-1.05) | 1.00<br>(0.92-1.09) | 1.0<br>(Ref) | 1.03<br>(0.94-1.12) | 0.98<br>(0.90-1.07) | 1.00<br>(0.92-1.09) | 1.04<br>(0.96-1.13) | 1.0<br>(Ref) |
| Leukemia | 1.02<br>(0.95-1.09) | 1.03<br>(0.97-1.09) | 1.01<br>(0.96-1.08) | 1.00<br>(0.94-1.06) | 1.0<br>(Ref) | 1.03<br>(0.97-1.10) | 1.03<br>(0.97-1.09) | 0.99<br>(0.94-1.05) | 1.03<br>(0.97-1.08) | 1.0<br>(Ref) |

Source: Adapted from Statistics Canada, Canadian Census Health and Environment Cohorts 2006 & 2011, Canadian Cancer Registry, 2006 long-form census, and 2011 National Household Survey.
CanCHEC = Canadian Census Health and Environment Cohort; 95% CI = confidence intervals computed using the Wald method; CNS = central nervous system; HL = Hodgkin's Lymphoma; NHL = Non-Hodgkin's Lymphoma.
Bolded age-standardized incidence rate ratio estimates are significant based on 95% confidence intervals.
<sup>a</sup> Model adjusted for 5-year age groups (except where indicated), education level (None, High school graduation certificate or equivalency certificate; Other trades certificate or diploma; Registered apprenticeship certificate; College, CEGEP or other non-university certificate or diploma from a program of 3 months to less than 1 year; College, CEGEP or other non-university certificate or diploma from a program of 1 year to 2 years; College, CEGEP or other non-university certificate or diploma from a program of more than 2 years; University certificate or diploma below bachelor level, Bachelor's degree; University certificate or diploma above bachelor level; Degree in medicine, dentistry, veterinary medicine, or optometry; Master's degree; Earned doctorate degree; Not applicable (institutional residents)), urbanicity (rural, urban) occupation industry sector (see Supplementary Table 2 for categories), province, and race.
<sup>b</sup> Age categories were collapsed for two cancer sites to assist model convergence. Breast cancer incidence rates were adjusted using ages 0-14, and 15-24, and 5-year age groups above 24. Multiple myeloma incidence rates were adjusted using age groups 0-29, and 10-year age groups above 29 (e.g., 30-39 years).

## DISCUSSION

This study estimated the associations between individual- and area-level income, and site-specific cancer incidence using a representative sample of the Canadian population. Given the coverage and size of this study population, our findings are generalizable to the entire Canadian population between 2006 and 2015 for all provinces except for Quebec after 2010. To our knowledge, this is the first study estimating site-specific cancer incidence by both individual- and area-level income quintile in Canada. The reported estimates of age-standardized and adjusted incidence rates suggest that one’s personal and neighbourhood wealth influence cancer incidence through distinct mechanisms.

Differences in site-specific cancer incidence across the individual-level income gradient point towards the potential differential distribution of lifestyle and environmental risk factors that affect cancer risk. Evidently, the observed trends in cancer incidence represent the weighted average of the effect of known and unknown risk factors that mediate the relationship between income and cancer incidence. Incidence was significantly greater among individuals of poorer income for cancer sites known to be associated with smoking, diet, and body weight (head and neck, esophageal, stomach, colorectal, liver, lung, and kidney and renal pelvis) (33). These established risk factors are more prevalent among poorer household income quintiles compared to the wealthiest in Canada (34). Infectious agents could contribute to decreasing incidence trends across the income gradient for stomach and liver cancer. In Canada, *Helicobacter pylori* positivity (35) as well as Hepatitis B and C seroprevalence (36) are higher among individuals of lower income. Changes in incidence rate ratios across the income gradient following adjustment indicate that factors adjusted for in our analyses (e.g., race, occupation, education) are also important drivers of risk, highlighting a complex interplay between elements of SES.

Disparities in utilization of cancer screening services by income could have influenced the observed incidence rates of colorectal, cervical, and breast cancer. In 2008, fewer Canadian individuals in lower household income quintiles were up to date for recommended cervical, breast, and colorectal cancer screening compared to those in wealthier household income quintiles, despite these screening services being covered by public health insurance (34). Cervical and colorectal screening aim to identify pre-cancerous conditions, whereas breast cancer screening aims to identify pre-existing tumours at an earlier stage. Higher colorectal and cervical cancer screening rates are therefore expected to lead to lower site-specific incidence, while higher breast cancer screening rates are expected to lead to more frequent diagnosis of breast cancer, and thus higher incidence. While these established disparities in screening attendance likely contributed to the observed trends, other risk factors, such as higher parity, which is protective against breast cancer (37) and increases the risk of cervical cancer (38), could have further driven the opposing incidence trends across individual-level income for both sites.

Even though our multivariable model was adjusted for race, a strong predictor of melanoma incidence (39), melanoma incidence persisted in being lower among poorer income groups. Diagnosis of melanoma is dependent upon access to dermatologic specialists. The probability of referral to specialists in Canada tends to be lower among individuals of poorer personal income despite more frequent visits to primary care providers (40). As intermittent sun exposure is a more important risk factor for melanoma than chronic sun exposure, it is plausible that those who are wealthier may have more intermittent sun exposure than those with less household income due to a higher prevalence of white collar occupations, which we only were partly able to adjust for, and also may be more likely to vacation in sunnier climates, both of which are known risk factors for melanoma (41, 42).

For uterine cancer (which included endometrial), we were expecting to see a higher incidence rate in lower income quintiles due to the higher prevalence of obesity in lower income quintiles (43). However, adjustment revealed little effect of area-level income on incidence, and the presence of a non-linear trend in incidence across individual-level income quintiles. It is likely that hysterectomy rates in Canada, which are higher among women of lower income (Supplemental Figure 1), influenced the observed trends. Although our adjusted IRRs reflect the true uterine cancer incidence by income group in Canada over the duration of follow up, they do not account for uterine cancer incidence among women at risk, or women with uteri. As our incidence rates were not hysterectomy-corrected, we may have underestimated the association between income and uterine cancer incidence among women at risk.

Area-level income likely has independent effects from individual-level income on cancer risk because it captures elements of the quality and type of built and social environment that individuals interact with (44). Built environment, or the man-made structuring of one’s physical surroundings, can influence an individual’s risk of cancer both directly and indirectly. Environmental exposures resulting from one’s built environment, such as water quality, ultraviolet radiation exposure, and air pollution, have direct effects on the development of cancer-related outcomes (45). Physical aspects of the environment, like neighbourhood walkability, greenspace, housing quality, public services, and food accessibility indirectly shape an individual’s health-related behaviours. An individual’s orientation within their physical surroundings modifies their health-promoting or -demoting behaviours, such as dietary, substance use, physical activity, and healthcare utilization choices (45). Policies on zoning and housing affordability impact the diversity, density, and homogeneity of a neighbourhood, which can change with whom and how an individual socially engages. In turn, an individual’s social environment molds their health-related beliefs, values, and choices (46). Collectively, variation in how an individual’s environment and social context are structured influence their lifestyle decisions, which can have direct effects on their health.

While there is overlap in the risk-based behaviours that mediate the effect of area- and individual-level income on cancer incidence, the mediating mechanisms function differently through both measurements of income. For instance, an individual’s personal income affects their ability to purchase high-quality food, however, their food purchasing is dependent upon the quality of options available in their environment. Other causal exposures that vary by neighbourhood wealth, such as air pollution (47), could directly impact an individual’s risk of cancer regardless of their personal income. Accordingly, the two measures of income together contextualize an individual’s health and behaviour and their feasibility to make health-related choices. The highly similar, but differing magnitudes of our estimated effects of individual- and area-level income on site-specific cancer incidence emphasize these distinct pathways through which personal and neighbourhood wealth affect cancer risk.

An important limitation to our findings was our inability to account for changes in individual household income and upward or downward social mobility over time. It is possible that income during critical life periods is more important in determining cancer risk than income within a few years from cancer diagnosis, which is what we measured in this study. Over an individual’s life course, there may be changes in personal and neighbourhood wealth. As the induction period for most cancers can range from years to decades, the most accurate measurement of the association between income and cancer incidence later in life would also include measurement of early-life socioeconomic exposures. Even though we were able to account for the time-varying nature of area-level income for the duration of follow-up in our analyses, we were unable to account for this allostatic load of socioeconomic exposure over the life course, nor exposure during potential critical life periods. Additionally, there may have been misclassification of individual-level income quintile for some older individuals after retirement. While the household after-tax income measure includes regular income from retirement pension plans, income funds, annuities, and old-age security, it does not count lump sum withdrawals made from retirement savings plans to supplement regular income (29). These lump sum withdrawals are more likely to occur between retirement and age 71, as after this age retirement savings plans are converted into an annuity or a registered retirement income fund, which are counted in household income. Nonetheless, such measurement error of ordinal exposures such as income biases estimates towards the null (1). Any bias due to exposure measurement error therefore likely led to an underestimation of the associations between of individual- and area-level income, and cancer incidence.

The reduction of inequalities in the incidence of cancer and other chronic conditions (48–50) requires more epidemiological research on the social determinants of health. Our findings suggest that area-level income influences cancer risk through mechanisms independent of individual-level income. Future investigation is needed to dissect the mediating risk factors through which the built and social environments influence cancer risk and how these are differentially distributed across the income gradient. Such research could aid in identifying modifiable risk factors linked to socioeconomic status and ultimately inform policymaking that maximizes equitable cancer prevention.

## Data Availability

Statistics Canada is the owner and steward of the data used in this report, and access to the data is regulated by the 1985 Statistics Act. To access the data, researchers must become deemed employees of Statistics Canada and sign a research contract. Members of post-secondary institutions such as a faculty, students, or staff may apply for data access to Statistics Canada microdata through the Research Data Centre program using the Microdata Access Portal (https://www.statcan.gc.ca/en/microdata/data-centres/access). Code used for the current analyses is available at the Borealis repository, at https://doi.org/10.5683/SP3/4Y15SN.

## Abbreviations

ASIR: Age-standardized incidence rates
ASIRR: Age-standardized rate ratios
CanCHEC: Canadian Census Health and Environment Cohort
CCR: Canadian Cancer Registry
CDA: Census dissemination area
CNS: Central nervous system
CVSD: Canadian Vital Statistics Death Registry
DRD: Derived record depository
HL: Hodgkin’s lymphoma
IRR: Incidence rate ratio
NHL: Non-Hodgkin’s lymphoma
NHS: National Household Survey
PCCF+: Postal Code^OM^ Conversion File Plus, Version D
SEER: Surveillance, Epidemiology, and End Results
SES: Socioeconomic status
T1PMF: T1 Personal Master File

## Data availability statement

Statistics Canada is the owner and steward of the data used in this report, and access to the data is regulated by the 1985 *Statistics Act*. To access the data, researchers must become deemed employees of Statistics Canada and sign a research contract. Members of post-secondary institutions such as a faculty, students, or staff may apply for data access to Statistics Canada microdata through the Research Data Centre program using the Microdata Access Portal (https://www.statcan.gc.ca/en/microdata/data-centres/access). Code used for the current analyses is available at the Borealis repository, at https://doi.org/10.5683/SP3/4Y15SN.

## Funding

This study was funded by a Canadian Institutes of Health Research (CIHR) (grant 179901) to EF, and by a CIHR HIV/AIDS and STBBI Research Initiative, sponsored by the CIHR Institute of Infection and Immunity, Postdoctoral Fellowship Award (support to SM; Funding Reference Number: 202110HIV-477526-93701).

## Conflict of Interest

TM and SM have no conflicts of interest to declare. PT received an MSc stipend from the Gerald Bronfman Department of Oncology, McGill University. ELF reports support for the present manuscript in the form of a grant to his institution in his name from the Canadian Institutes of Health Research and the Cancer Research Society; consultancy for Merck; and financial interests with Elsevier and Elifesciences Ltd in the form of support fees to maintain the editorial office and work as Senior Editor, respectively. MZ and ELF hold a patent related to the discovery “DNA methylation markers for early detection of cervical cancer”, registered at the Office of Innovation and Partnerships, McGill University, Montreal, Quebec, Canada.

## Acknowledgements

Data Source: Statistics Canada, Canadian Census Health and Environment Cohorts 2006 & 2011, *2006 long-form census*, 2011 National Household Survey, Canadian Vital Statistics Death Database 2006-2015, and Canadian Cancer Registry 2006-2015. The Postal Code^OM^ Conversion File Plus (7D) is based on data licensed from Canada Post Corporation. Reproduced and distributed on an “as is” basis with the permission of Statistics Canada. This does not constitute an endorsement by Statistics Canada of this product.

## Notes

### Author Declarations

Ethics committee/IRB of McGill University gave ethical approval for this work.

