## Supplemental Tables 1-2, Supplemental Figure 1 for "Differences in site-specific cancer incidence by individual- and area-level income in Canada from 2006-2015"

**SUPPLEMENT**

### Table of Contents

**Supplementary Table 1. ICD-O-3 Topography and Histology codes by cancer site based on SEER site groupings. 1**

**Supplementary Table 2. Distribution of individual-level and area-level income quintiles by industry sector for the 2006 and 2011 CanCHECs. 2**

**Supplementary Figure 1. Proportion of female respondents of 2012 Canadian Community Health Survey respondents greater than 18 years of age who have had a hysterectomy, by 5-year age group. Adapted from the 2012 Canadian Community Health Survey. 4**

**Supplementary Table 1.** ICD-O-3 Topography and Histology codes by cancer site based on SEER site groupings

| Cancer site | ICD-O-3 Topography | ICD-O-3 Histology (Type) |
|---|---|---|
| All cancers | All sites C00-C80 | All invasive sites |
| Head and neck[a] | C00-C14, C300-C329 | 8000-9049, 9056-9139,9141-9589 |
| Esophagus | C150-C159 | 8000-9049, 9056-9139,9141-9589 |
| Stomach | C160-C169 | 8000-9049, 9056-9139,9141-9589 |
| Colorectal | C180-C189, C260, C199, C209 | 8000-9049, 9056-9139,9141-9589 |
| Anal | C210-C212, C218 | 8000-9049, 9056-9139,9141-9589 |
| Liver | C220 | 8000-9049, 9056-9139,9141-9589 |
| Pancreas | C250-C259 | 8000-9049, 9056-9139,9141-9589 |
| Lung & bronchus | C340-C349 | 8000-9049, 9056-9139,9141-9589 |
| Melanoma | C440-C449 | 8720-8790 |
| Breast | C500-C509 | 8000-9049, 9056-9139,9141-9589 |
| Cervix (female) | C530-C539 | 8000-9049, 9056-9139,9141-9589 |
| Uterus (female) | C540-C549, C559 | 8000-9049, 9056-9139,9141-9589 |
| Ovary (female) | C569 | 8000-9049, 9056-9139,9141-9589 |
| Prostate (male) | C619 | 8000-9049, 9056-9139,9141-9589 |
| Testis (male) | C620-C629 | 8000-9049, 9056-9139,9141-9589 |
| Bladder | C670-C679 | 8000-9049, 9056-9139,9141-9589 |
| Kidney & renal pelvis | C649, C659 | 8000-9049, 9056-9139,9141-9589 |
| Brain & CNS | C710-C719 | 8000-9049, 9056-9139,9141-9589 |
| | C710-C719 | 9530 - 9539 |
| | C700-C709, C720-C729 | 8000-9049, 9056-9139,9141-9589 |
| Thyroid | C739 | 8000-9049, 9056-9139,9141-9589 |
| Hodgkin Lymphoma | C000-C809 | 9650-9667 |
| Non-Hodgkin Lymphoma | C000-C809 | 9590-9597, 9670-9729, 9735-9738 |
| | All topographies excluding (C420, C421, C424) | 9811-9818, 9823, 9827, 9837 |
| Multiple myeloma | C000-C809 | 9731-9732, 9734 |
| Leukemia | C000 - C809 | 9826, 9835-9836 |
| | C420, C421, C424 | 9811-9818, 9837 |
| | C420, C421, C424 | 9840, 9861, 9865, 9866, 9867, 9869, 9871-9874, 9895-9897, 9898, 9910, 9911, 9920 |
| | C000 - C809 | 9863, 9875, 9876, 9945, 9946 |
| | C000 - C809 | 9733, 9742, 9800, 9801, 9805, 9806, 9807, 9808, 9809, 9820, 9831, 9832, 9833, 9834, 9860, 9870, 9891, 9930, 9931, 9940, 9948, 9963, 9964 |
| | C420, C421, C424 | 9827 |

Source: Canadian Cancer Statistics Advisory Committee, in collaboration with the Canadian Cancer Society, Statistics Canada, the Public Health Agency of Canada. Canadian Cancer Statistics 2021. Toronto, ON; 2021 (1).
[a] Definition based on the Canadian Cancer Statistics 2021 rather than SEER site groupings.

**Supplementary Table 2.** Distribution of individual-level and area-level income quintiles by industry sector for the 2006 and 2011 CanCHECs

| | 2006 CanCHEC | | | | | | | 2011 CanCHEC | | | | | | |
|---|---|---|---|---|---|---|---|---|---|---|---|---|---|---|
| | Individual-level income quintile | | | Area-level income quintile | | | | Individual-level income quintile | | | Area-level income quintile | | | |
| **Occupation (%)** | **Q1** | **Q3** | **Q5** | **Q1** | **Q3** | **Q5** | **NA[a]** | **Q1** | **Q3** | **Q5** | **Q1** | **Q3** | **Q5** | **NA[a]** |
| Agriculture, forestry, fishing and hunting | 3 | 2 | 1 | 2 | 2 | 2 | 5 | 2 | 2 | 1 | 1 | 2 | 1 | 2 |
| Mining and oil and gas extraction | <1 | 1 | 2 | 1 | 1 | 1 | 1 | <1 | 1 | 2 | 1 | 1 | 1 | 1 |
| Utilities | <1 | <1 | 1 | <1 | <1 | 1 | <1 | <1 | <1 | 1 | <1 | 1 | 1 | 1 |
| Construction | 3 | 4 | 4 | 3 | 4 | 3 | 4 | 3 | 4 | 4 | 3 | 4 | 3 | 4 |
| Manufacturing | 3 | 8 | 8 | 6 | 7 | 6 | 3 | 3 | 6 | 6 | 5 | 6 | 5 | 2 |
| Wholesale trade | 1 | 3 | 4 | 2 | 3 | 3 | 1 | 1 | 2 | 3 | 2 | 3 | 3 | 1 |
| Retail trade | 5 | 7 | 6 | 6 | 7 | 6 | 4 | 5 | 7 | 6 | 6 | 7 | 6 | 5 |
| Transportation and warehousing | 2 | 3 | 3 | 3 | 3 | 2 | 2 | 2 | 3 | 3 | 2 | 3 | 2 | 2 |
| Information and cultural industries | 1 | 1 | 2 | 1 | 1 | 2 | 1 | 1 | 1 | 2 | 1 | 1 | 2 | 1 |
| Finance and insurance | 1 | 2 | 4 | 1 | 2 | 3 | 1 | 1 | 2 | 5 | 2 | 3 | 3 | 1 |
| Real estate and rental and leasing | 1 | 1 | 2 | 1 | 1 | 1 | 1 | 1 | 1 | 1 | 1 | 1 | 1 | 1 |
| Professional, scientific and technical services | 2 | 3 | 7 | 2 | 4 | 6 | 2 | 2 | 4 | 8 | 3 | 4 | 6 | 2 |
| Management of companies and enterprises | <1 | <1 | <1 | <1 | <1 | <1 | <1 | <1 | <1 | <1 | <1 | <1 | <1 | <1 |
| Administrative and support, waste management and remediation services | 3 | 3 | 2 | 3 | 2 | 2 | 2 | 3 | 2 | 2 | 3 | 2 | 2 | 2 |
| Educational services | 2 | 4 | 7 | 3 | 4 | 6 | 5 | 2 | 4 | 8 | 4 | 5 | 6 | 4 |
| Health care and social assistance | 3 | 6 | 8 | 5 | 6 | 7 | 5 | 4 | 7 | 9 | 6 | 7 | 7 | 5 |
| Arts, entertainment and recreation | 1 | 1 | 1 | 1 | 1 | 2 | 1 | 1 | 1 | 1 | 1 | 1 | 2 | 1 |
| Accommodation and food services | 5 | 4 | 3 | 4 | 4 | 4 | 5 | 4 | 4 | 3 | 4 | 4 | 3 | 3 |

| | | | | | | | | | | | | | | |
|---|---|---|---|---|---|---|---|---|---|---|---|---|---|---|
| Other services (except public administration) | 2 | 3 | 3 | 3 | 3 | 3 | 2 | 2 | 3 | 3 | 2 | 3 | 2 | 2 |
| Public administration | 2 | 3 | 7 | 3 | 4 | 4 | 9 | 2 | 4 | 8 | 4 | 5 | 5 | 10 |
| Not reported [b] | 60 | 41 | 24 | 49 | 40 | 38 | 47 | 61 | 41 | 24 | 49 | 40 | 39 | 49 |

Source: Adapted from Statistics Canada, Canadian Census Health and Environment Cohorts 2006 & 2011, 2006 long-form census, and 2011 National Household Survey.
CanCHEC = Canadian Census Health and Environment Cohorts.
[a] NA indicates category of individuals for whom area-level income quintiles could not be assigned due to missing area-level median income data.
[b] Age < 15 years, worked before 2000, or never worked.

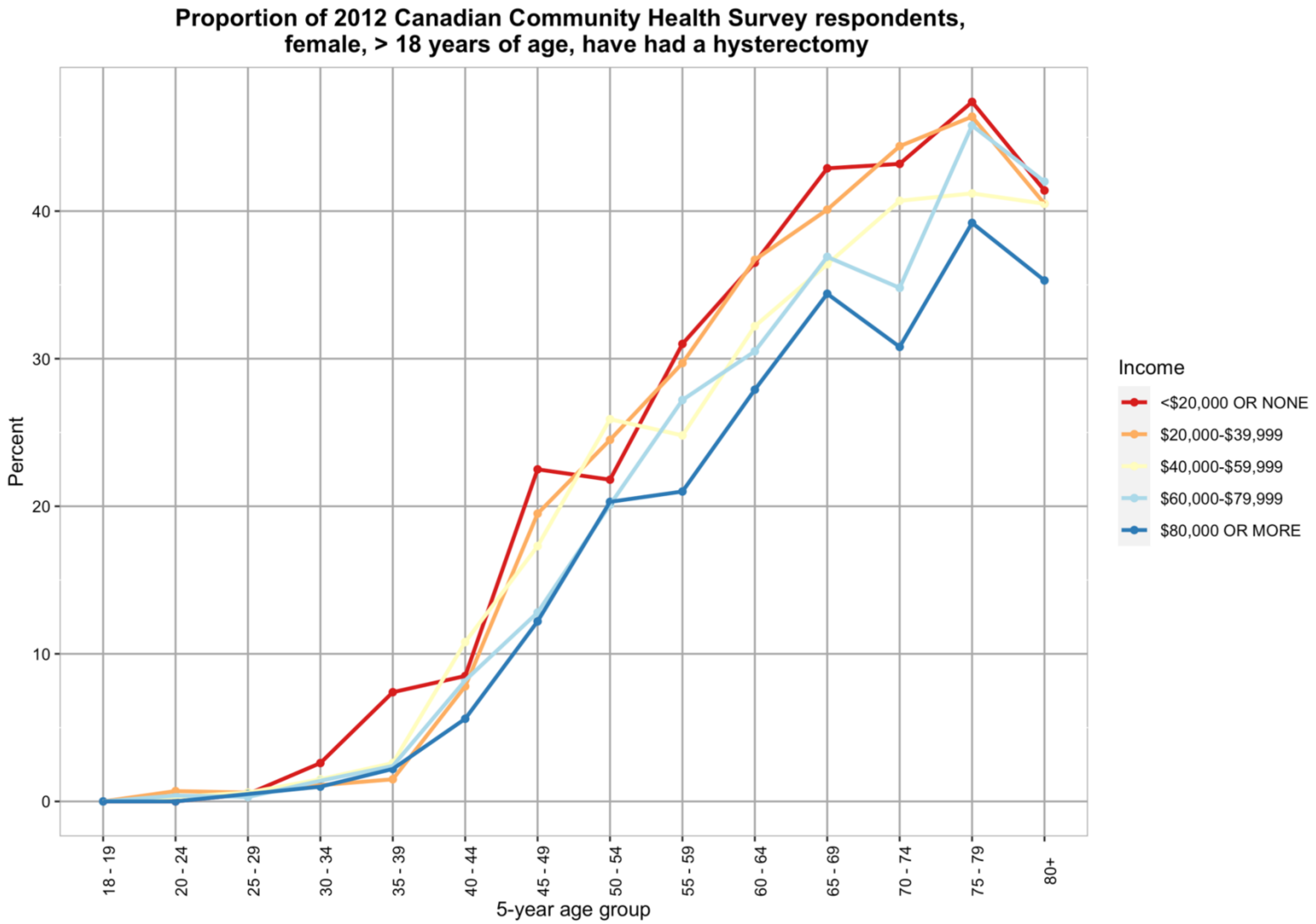


**Supplementary Figure 1.** Proportion of female respondents of 2012 Canadian Community Health Survey respondents greater than 18 years of age who have had a hysterectomy, by 5-year age group and household income. Adapted from the 2012 Canadian Community Health Survey (2).

Supplement References

1. Canadian Cancer Statistics Advisory Committee, in collaboration with the Canadian Cancer Society, Statistics Canada, the Public Health Agency of Canada. Canadian Cancer Statistics 2021. Toronto, ON; 2021.

2. Statistics Canada. Canadian Community Health Survey, 2012 2014 [Available from: https://www23.statcan.gc.ca/imdb-bmdi/instrument/3226_Q1_V9-eng.htm.
